# Impact of Social Determinants of Health in Early Pregnancy on Racial and Ethnic Differences in Post-pregnancy Cardiovascular Health

**DOI:** 10.64898/2026.09.14.26363034

**Authors:** Xiaoning Huang, Lucia C Petito, Lynn M Yee, Natalie Cameron, David M Haas, Brian M Mercer, Samuel Parry, George R Saade, Robert M Silver, Yi Qiao, Xiaomeng Huang, Hyagriv N Simhan, Uma M. Reddy, Judith Chung, Philip Greenland, Donald M Lloyd-Jones, Kiarri N Kershaw, William A Grobman, Sadiya S Khan, Kartik K Venkatesh

**Author notes:** **Correspondence to:** Xiaoning Huang, PhD,. Department of Medicine, Division of Cardiology, Northwestern University Feinberg School of Medicine, 680 N Lakeshore Dr, Suite 1400, Chicago, IL 60611.

## Abstract

**Background:** Social determinants of health (SDOH) influence maternal cardiovascular health (CVH), but the extent to which SDOH in early pregnancy may explain racial and ethnic differences in post-pregnancy CVH has not been defined.

**Methods:** This secondary analysis used data from the prospective multisite Nulliparous Pregnancy Outcomes Study: Monitoring Mothers-to-Be Heart Health Study. CVH was assessed in early pregnancy and 2-7 years after delivery using the American Heart Association’s Life’s Essential 8 (LE8) framework. We used the Oaxaca-Blinder decomposition to quantify the statistical contributions of differences in early pregnancy CVH, SDOH, psychosocial factors, and demographic covariates to racial and ethnic differences in post-pregnancy CVH between the two largest minoritized racial and ethnic groups (non-Hispanic [NH] Black and Hispanic) compared with NH White women.

**Results:** Among 4,088 women (14.4% Black, 17.6% Hispanic, and 68.0% White), the mean (SD) post-pregnancy LE8 score was lower in Black (70.1 [14.1]) and Hispanic (77.8 [12.7]) women compared with White women (82.4 [12.7]) (all p<0.01). Independent of early pregnancy CVH, maternal age, and nativity, differences in SDOH were associated with 4.5 (95% CI, 2.8-6.2) points of the Black-White gap (37% of the observed post-pregnancy CVH difference) and 3.2 (95% CI, 1.1-5.3) points of the Hispanic-White gap (70% of the observed post-pregnancy CVH difference), respectively.

**Conclusions:** Most racial and ethnic differences in post-pregnancy CVH 2 to 7 years after a first birth were explained by differences in early pregnancy CVH and SDOH. Independent of early pregnancy CVH, SDOH remained the largest measured contributor to the residual difference.

**CLINICAL PERSPECTIVE:** *What Is New?:* Among 4,088 women followed 2 to 7 years after a first birth, differences in early pregnancy social determinants of health and cardiovascular health together accounted for approximately three-quarters of the racial and ethnic differences in post-pregnancy cardiovascular health.

*What Are the Clinical Implications?:* Because differences in cardiovascular health persisted beyond what differences in measured social risk factors explained, social risk screening in early pregnancy should be paired with sustained cardiovascular follow-up after delivery rather than treated as sufficient on its own. Black women did not experience the improvements in physical activity and smoking cessation seen in other groups after a first birth, identifying the postpartum period as a window for behavioral cardiovascular risk reduction.

## INTRODUCTION

The peripartum period is a critical window for assessing maternal cardiovascular health (CVH) and modifying long-term cardiovascular risk.^1^ Poor CVH in early pregnancy is associated with adverse maternal and neonatal outcomes and severe maternal morbidity and mortality.^2–5^ In addition, poor CVH is associated with cardiovascular disease (CVD) and long-term adverse cardiometabolic outcomes postpartum for both the mother and offspring via fetal programming.^6^ Recent data from the United States (US) demonstrate that CVH in the peripartum period has worsened over the past two decades, and more so for minoritized racial and ethnic women.^7,8^

The American Heart Association (AHA) defines composite CVH using eight health factors and behaviors collectively termed Life’s Essential 8 (LE8), which includes body mass index (BMI), blood pressure, blood cholesterol, blood glucose, dietary quality, physical activity, sleep health, and tobacco use.^9^ LE8 is a validated measure associated with an increased risk of cardiovascular disease (CVD).^9^ Professional societies have identified optimizing peripartum CVH as a key strategy for improving maternal health.^4^

Persistent and significant racial and ethnic disparities in the peripartum period contribute to adverse pregnancy and cardiometabolic outcomes.^10–12^ Minoritized subgroups, including non-Hispanic Black and Hispanic women, experience lower or worse LE8 scores in early pregnancy compared with non-Hispanic White women.^13^ The 2024 AHA/ACC Joint Report provides a standardized framework for identifying and measuring Social Determinants of Health (SDOH) to reduce CVH disparities.^14^ Empirical research have shown that individual-and neighborhood-level social determinants of health (SDOH), including socioeconomic status, health literacy, access to care, psychosocial well-being, perceived discrimination, and environmental factors, contribute to racial and ethnic disparities in maternal CVH.^15–17^

Prior studies have focused on identifying associations between SDOH and peripartum CVH by race and ethnicity; however, a decomposition framework can quantify the expected change in CVH between racial and ethnic groups if SDOH factors were equalized across racial and ethnic groups.^18^ Characterizing risk factors in early pregnancy for long-term maternal CVH disparities can inform public health interventions and policies that address the root causes of inequity across a woman’s life course.^19^ For example, a recent decomposition analysis found that SDOH factors explained over 80% of racial and ethnic disparities in early pregnancy CVH,^13^ but the extent to which SDOH and CVH in early pregnancy continue to explain persistent racial and ethnic differences in long-term CVH after delivery remains unknown.

Pregnancy functions as a physiologic stress test, and the social conditions in which a pregnancy occurs shape how a woman meets it. Nutrition insecurity, limited access to care, chronic stress, and disadvantaged neighborhood environments are experienced disproportionately by minoritized racial and ethnic groups, and the metabolic and hemodynamic adaptations of pregnancy may not fully resolve when those conditions persist after delivery. Early pregnancy is also, for many women, the first sustained contact with the health care system, which makes it an actionable point of intervention regardless of the underlying mechanism. Characterizing the social conditions present at that moment is therefore relevant both to understanding why differences in CVH persist and to identifying when they might be addressed.

We examined the extent to which differences in early pregnancy SDOH, spanning individual socioeconomic, neighborhood, and social context domains, together with psychosocial factors and early pregnancy CVH, account for racial and ethnic differences in CVH 2-7 years after delivery.

## METHODS

The data that support the findings of this study are available from the Eunice Kennedy Shriver National Institute of Child Health and Human Development Data and Specimen Hub (DASH) for investigators who meet the criteria for access to de-identified data. Analytic code used to generate the results reported here is available from the corresponding author upon reasonable request. No new materials were generated for this analysis.

### Study Sample

This is a secondary analysis of data from the multicenter, prospective Nulliparous Pregnancy Outcomes Study: Monitoring Mothers-to-Be (nuMoM2b-HHS) cohort. The nuMoM2b study enrolled nulliparous individuals with singleton pregnancies with cardiac activity at less than 14 weeks of gestation between September 2010 and June 2014 across eight clinical sites affiliated with tertiary care medical centers in the US.^20^ The nuMoM2b-HHS enrolled a representative sub-cohort of nuMoM2b participants to extend follow-up through 2 to 7 years after delivery to evaluate CVH after a first birth.^21^ Institutional review board approval and written informed consent were obtained at each participating institution.^20^ Study identifiers for 8,838 nuMoM2b participants who had delivery data and consented to future contact were released for interval contacts conducted every 6 to 12 months. Of these, 7,872 were reached, and 867 declined to participate further. Among the 7,003 who completed one or more interval contacts, 6,576 were eligible for an in-person visit during at least one wave, 5,206 agreed to a visit, and 4,508 attended. Attendance required that a participant was not currently pregnant, confirmed by a pregnancy test administered at the start of the visit, and was more than 6 months postpartum from any subsequent pregnancy. The follow-up assessment was an in-person research visit during which height, weight, and blood pressure were measured by trained personnel, a fasting blood sample was obtained, and participants completed interviewer-administered and self-administered questionnaires.

Of 4,508 participants in the nuMoM2b-HHS cohort, the current analysis included 4,088 (92.3%) adult participants who self-identified as Hispanic, Non-Hispanic Black (Black, hereafter), or Non-Hispanic White (White, hereafter). We excluded individuals identifying as other racial and ethnic groups (e.g., non-Hispanic Asian and multiracial) due to insufficient sample size to support decomposition analyses. Race and ethnicity were self-reported at study enrollment, treated as social constructs, and defined to be consistent with NCHS Vital Statistics criteria.^22^ Characteristics of participants in the analytic sample and of those not included are compared in Supplemental Table S1. Participation in the follow-up visit was associated with measured baseline characteristics, which are adjusted for in all models; analyses assume that participation is independent of post-pregnancy CVH conditional on those measured covariates.

### CVH Measurements

CVH using the LE8 framework was assessed in early pregnancy at a mean gestational age of 11.4 weeks and maternal age of 27.1, and then again at the follow-up visit 2-7 years after delivery. CVH was measured using the composite LE8 CVH score, quantified by eight clinical and behavioral measures: BMI, blood pressure, non-HDL cholesterol, fasting glucose, dietary quality (Healthy Eating Index 2010),^23^ moderate-to-vigorous physical activity, sleep duration, and current smoking status.^9^ Briefly, each component received a score of 0 to 100, with 0 representing the least favorable and 100 representing the most favorable CVH. The composite LE8 CVH score was calculated as the individual-specific mean of the scores of the eight components, yielding a total score ranging from 0 to 100. Height, weight, and blood pressure were measured by trained study personnel using standardized protocols.^20^ Early pregnancy lipid and glucose values were obtained from non-fasting blood samples while follow-up measures were obtained from fasting samples. Data on dietary quality, physical activity, sleep duration, and tobacco use were obtained via validated self-report instruments recommended for the LE8 scoring algorithm.^9^ Dietary quality was assessed with the Block/Bodnar/nuMoM2b 2010 Food Frequency Questionnaire, a modification of the Block 2005 Food Frequency Questionnaire comprising approximately 120 food and beverage items, self-administered in approximately 20 to 30 minutes and analyzed by NutritionQuest.^20,24^ The same instrument was administered at both time points, with a reference period of the 3 months before conception in early pregnancy and the past year at follow-up; the early pregnancy dietary measure is therefore a pre-pregnancy measure.^21^ Physical activity was assessed with the Modifiable Activity Questionnaire,^25^ from which leisure time moderate-to-vigorous physical activity was derived using metabolic equivalent values from the 2011 Compendium of Physical Activities,^26^ excluding occupational and household activities and activities below 3.0 METs, with vigorous intensity minutes weighted twice.^27^ Sleep was assessed as self-reported average hours of sleep per night. Tobacco use was assessed by self-reported smoking status.

LE8 component scores were assigned using the thresholds specified by the AHA presidential advisory for blood pressure, body mass index, non-HDL cholesterol, physical activity, and sleep duration.^9^ Operationalization details were listed in Supplemental Table S5.

### Social Determinants of Health

We organized early pregnancy SDOH using the 2024 ACC/AHA Key Data Elements and Definitions for Social Determinants of Health in Cardiology as an organizing framework, which distinguishes individual socioeconomic circumstances, neighborhood and built environment, health care access, and social context.^14^ Perceived racial discrimination, a social context measure, was grouped with individual-level SDOH for estimation. Psychosocial factors, including resilience, perceived social support, anxiety and depressive symptoms, and perceived stress, are established correlates of CVH but are also hypothesized to lie on the pathway between structural social determinants and CVH.^28^ Therefore, we modeled them as a distinct block and report the contribution of SDOH both with and without adjustment for them.

Demographic factors included: age at the first study visit and nativity (US-born vs. non-US-born). Individual-level SDOH factors included: educational attainment (years of schooling), total annual family income (in $10,000 USD), health insurance type (private, public, or other), and health literacy per the Rapid Estimate of Adult Literacy in Medicine Short Form, dichotomized per adequacy (score ≥7 vs. <7).^24^ Perceived discrimination was measured by whether participants reported ≥ 3 lifetime experiences of racial discrimination.^29^ Neighborhood-level socioeconomic status was measured by the Area Deprivation Index (ADI), mapping each census block group to a national rank (0 as most advantaged to 100 as most disadvantaged).^30^ Psychosocial factors included resilience per the Connor-Davidson Resilience Scale (range 0-100),^31^ perceived social support per the Multidimensional Scale of Perceived Social Support (range 0-72),^32^ anxiety symptoms per the State-Trait Anxiety Inventory (range 0-60),^33^ depressive symptoms per the Edinburgh Postnatal Depression Scale (range 0-30),^34^ and perceived stress per the Perceived Stress Scale-10 (range 0-40).^35^ Nativity and SDOH data were collected via structured interview and psychosocial factors were self-reported via standardized survey.^20^ Measures were obtained at the first study visit (6 to 13 weeks of gestation) with the following exceptions, which were administered at the second study visit (16 to 21 weeks of gestation) and are used here as proxies to early pregnancy measures: health literacy, perceived racial discrimination, resilience, perceived social support, and anxiety symptoms.

### Statistical Analysis

Descriptive statistics were computed for the overall sample and stratified by self-identified race and ethnicity. The Oaxaca-Blinder decomposition method was used to quantify the contributions of early pregnancy CVH and SDOH factors to racial and ethnic differences in CVH 2-7 years after pregnancy.^36,37^ This statistical approach decomposes the mean difference in post-pregnancy CVH scores between racial and ethnic groups (the post-pregnancy CVH gap) into two components: an *explained* component that represents the portion of the post-pregnancy CVH gap attributable to differences in early pregnancy CVH and SDOH factors between groups; and an *unexplained* component (also termed the “effect modification” component), which represents the portion of the post-pregnancy CVH gap attributable to differences in the associations (i.e., regression coefficients) between early pregnancy factors and post-pregnancy CVH between racial and ethnic groups.^38^

The *explained* component quantifies the expected reduction in the gap in post-pregnancy CVH between Black or Hispanic women and White women if early pregnancy CVH and SDOH of Black and Hispanic women were the same as those of White women; these analyses retain the original race-and ethnicity-specific associations (regression coefficients) between early pregnancy CVH and post-pregnancy CVH. The *unexplained* component quantifies the expected reduction in the gap in post-pregnancy CVH between racial and ethnic groups if the magnitude of the associations between SDOH and early pregnancy CVH and post-pregnancy CVH among Black or Hispanic women were the same as those observed among White women; these analyses retain the original levels of SDOH and early pregnancy CVH. All analyses assume there are no other factors associated with differences in post-pregnancy CVH scores that operate through paths excluding all measured SDOH and early pregnancy CVH. White women were defined as the reference racial and ethnic group because they had the most favorable CVH and were the majority of the sample.^13^ Multiple Imputation by Chained Equations was used to impute missing data with 10 complete datasets.^39^ All analyses were performed in each dataset and results were combined using Rubin’s rules.^40^ All analyses were performed in Stata 18 (StataCorp LLC, College Station, TX), two-fold decomposition via the “oaxaca” command with linear regression^41^ and multiple imputation by chained equations via the “mi chained” command^39^ (missingness and models used for imputation were included in **Supplemental Table S2**). To address the identification problem inherent in the decomposition of categorical variables, the normalized option was used, which averaged estimates obtained from varying the reference group.^42^ Standard errors were clustered around nuMoM2b study sites. Statistical significance was interpreted at p<0.05. For each comparison, post-pregnancy LE8 score was regressed on the full vector of early pregnancy covariates in a linear model fit within each racial and ethnic group, and the two-fold decomposition was computed using the pooled coefficients as the reference structure. All covariates entered a single equation simultaneously; the blocks reported in the tables are groupings of the detailed contributions for presentation and do not represent sequential entry into the model.

To address the possibility that psychosocial factors lie on the causal pathway between SDOH and CVH, we estimated the decomposition under three model specifications. Model 1 included demographic covariates and SDOH. Model 2 added psychosocial factors. Model 3, the primary model, added early pregnancy CVH. Comparison of Models 1 and 2 quantifies how much of the contribution of SDOH operates through measured psychosocial factors. Model 3 is not only a more conservative version of the estimate but also quantifies the contribution of SDOH to differences in post-pregnancy CVH that does not operate through CVH already present in early pregnancy, which is the quantity relevant to what addressing SDOH could add beyond optimization of early pregnancy CVH.

Three supplementary analyses were performed. First, the decomposition was repeated among participants with complete data on all analytic variables. Second, model assumptions were examined by plotting residuals against fitted values and against normal quantiles within each racial and ethnic group. Third, to characterize how CVH changed between early pregnancy and follow-up, the within-group mean difference in each LE8 component score was estimated with 95% confidence intervals; these within-group changes are descriptive and were not compared across groups.

## RESULTS

### Sample characteristics and racial and ethnic differences in CVH

Of the 4,088 included participants, 2,778 (68.0%) identified as White, 590 (14.4%) as Black, and 720 (17.6%) as Hispanic. Individual-and neighborhood-level SDOH measured in early pregnancy varied across racial and ethnic groups (**Table 1**). Black (mean age, 24.0 [SD, 5.2] years) and Hispanic (25.1 [5.3]) women were younger at enrollment compared with White women (28.3 [5.1]). Black and Hispanic women had lower levels of educational attainment, annual household income, and health literacy, and higher public health insurance enrollment compared with White women. More Hispanic women were born outside the US compared with Black and White women. Neighborhood-level socioeconomic disadvantage as measured by the ADI was higher for Black and Hispanic women compared with White women. Black women reported more anxiety symptoms, depressive symptoms, and perceived stress and less social support compared with White women. Early pregnancy CVH as measured by LE8 score (mean [SD]) was worse (i.e., lower scores) for Black (72.4 [12.9]) and Hispanic women (77.6 [11.8]) compared with White women (82.4 [12.9]).

**Table 1.** Early pregnancy Demographic, SDOH, and Psychosocial Characteristics, and LE8 Overall and by Racial and Ethnic Group (nuMoM2b-HHS, N=4,088)

|  | **Total**<br>**N=4,088** | **NH White**<br>**N=2,778** | **NH Black**<br>**N=590** | **Hispanic**<br>**N=720** |
| --- | --- | --- | --- | --- |
| **Demographic characteristics** |  |  |  |  |
| Maternal Age, years, mean (SD) | 27.1 (5.4) | 28.3 (5.1) | 24.0 (5.2) | 25.1 (5.3) |
| Nativity |  |  |  |  |
| US-born, % | 88.9% | 96.0% | 94.6% | 56.8% |
| Born outside the US, % | 11.1% | 4.0% | 5.4% | 43.2% |
| Gestational age in weeks at enrollment, mean (SD) | 11.4 (1.6) | 11.4 (1.6) | 11.5 (1.6) | 11.5 (1.5) |
| **Individual SDOH characteristics** |  |  |  |  |
| Education (years of schooling), mean (SD) | 15.2 (2.4) | 15.9 (2.2) | 13.5 (2.2) | 14.2 (2.4) |
| Household income (per \$10,000), mean (SD) | 7.6 (5.8) | 9.0 (5.7) | 3.7 (4.0) | 5.0 (4.7) |
| Health insurance |  |  |  |  |
| Others, % | 3.3% | 2.9% | 4.4% | 4.2% |
| Covered by public insurance, % | 26.7% | 12.5% | 58.9% | 55.3% |
| Covered by private insurance, % | 70.0% | 84.7% | 36.7% | 40.5% |
| Adequate health literacy, % | 82.6% | 92.6% | 64.6% | 58.8% |
| **Neighborhood-SDOH characteristics** |  |  |  |  |
| Area deprivation index 2015, national rank, mean (SD) | 48.1 (30.5) | 39.2 (25.6) | 74.4 (25.4) | 61.0 (34.1) |
| **Social context** |  |  |  |  |
| Experienced racial discrimination, % | 5.9% | 1.9% | 18.8% | 10.8% |
| **Psychosocial factors** |  |  |  |  |
| Connor-Davidson Resilience Scale (0-100), mean (SD) | 79.0 (12.2) | 79.6 (11.3) | 78.7 (14.3) | 77.0 (13.6) |
| Multidimensional Scale of Perceived Social Support (0-72), mean (SD) | 62.3 (14.1) | 63.5 (13.4) | 57.8 (16.0) | 61.2 (13.9) |
| State-Trait Anxiety Inventory (0-60), mean (SD) | 13.8 (8.8) | 13.4 (8.6) | 15.0 (9.2) | 14.5 (9.1) |
| Edinburgh Postnatal Depression Scale (0-30), mean (SD) | 5.7 (4.2) | 5.4 (4.0) | 6.4 (4.8) | 6.2 (4.4) |
| Perceived Stress Scale-10 (0-40), mean (SD) | 12.6 (6.7) | 11.8 (6.3) | 15.0 (7.3) | 13.6 (7.0) |
| LE8 CVH Score, mean (SD) | 80.1 (13.2) | 82.4 (12.9) | 72.4 (12.9) | 77.6 (11.8) |

Data on means and percentages are combined from 10 imputed datasets using Rubin's rules. Abbreviations. LE8: Life's Essential 8; CVH: cardiovascular health; NH: Non-Hispanic; SD: standard deviation

CVH 2-7 years after delivery as measured by LE8 score (mean [SD]) was lower for Black (70.1 [14.1]) and Hispanic women (77.8 [12.7]) compared with White women (82.4 [12.7]) (**Table 2**). Racial and ethnic differences were observed in LE8 component scores as well. Black women had the lowest scores for blood pressure, fasting glucose, BMI, diet, physical activity, sleep health, and tobacco use among the three groups. Hispanic women generally had lower scores for most LE8 components compared with White women except for blood pressure and non-HDL cholesterol. White women had the lowest score for non-HDL cholesterol compared with Black and Hispanic women. Early pregnancy and post-pregnancy data were similar across both imputed and complete case data (**Supplementary Table S3 & S4**).

**Table 2.** Post-pregnancy Cardiovascular Health Factors and Composite LE8 CVH Score 2 to 7 Years After Delivery, Overall and by Racial and Ethnic Group (nuMoM2b-HHS, N=4,088)

|  | **Total**<br>**N=4,088** | **NH White**<br>**N=2,778** | **NH Black**<br>**N=590** | **Hispanic**<br>**N=720** |
| --- | --- | --- | --- | --- |
| **Life's Essential 8 Scores, mean (SD)** |  |  |  |  |
| **Blood Pressure Score** | 85.8 (24.8) | 86.8 (23.5) | 79.3 (29.6) | 87.2 (24.1) |
| **Lipid Score** | 78.7 (27.4) | 77.5 (28.0) | 84.4 (24.4) | 78.8 (26.8) |
| **Glucose Score** | 91.1 (22.5) | 92.4 (20.5) | 86.9 (28.3) | 89.6 (24.0) |
| **BMI Score** | 67.5 (35.8) | 72.8 (33.7) | 48.0 (38.2) | 63.3 (35.3) |
| **Diet Score** | 70.4 (25.1) | 73.7 (23.0) | 55.0 (29.6) | 70.3 (24.2) |
| **Physical Activity Score** | 68.0 (40.7) | 74.4 (37.4) | 52.7 (43.9) | 55.9 (44.3) |
| **Sleep Health Score** | 87.3 (20.5) | 89.2 (18.8) | 80.8 (24.3) | 85.5 (22.0) |
| **Nicotine Exposure Score** | 89.4 (30.7) | 92.2 (26.8) | 73.9 (43.9) | 91.5 (27.9) |
| **Overall CVH** | 79.8 (13.6) | 82.4 (12.7) | 70.1 (14.1) | 77.8 (12.7) |
| **CVH Factors, mean (SD)** |  |  |  |  |
| **Mean Arterial Pressure (mmHg)** | 85.3 (9.8) | 85.0 (9.4) | 87.4 (11.5) | 84.8 (9.3) |
| **Non-HDL-cholesterol (mg/dL)** | 126.1 (37.5) | 128.4 (38.1) | 116.0 (35.2) | 125.6 (35.5) |
| **Glucose (mg/dL)** | 90.9 (20.9) | 89.7 (17.3) | 95.1 (34.8) | 92.1 (18.2) |
| **BMI (kg/m<sup>2</sup>)** | 27.9 (7.7) | 26.7 (7.0) | 32.2 (9.2) | 28.8 (7.4) |
| **HEI-2010 Total Score** | 62.1 (12.0) | 63.8 (11.5) | 54.8 (11.8) | 61.8 (11.6) |
| **Physical activity (min/week)** | 249.0 (279.3) | 274.9 (278.9) | 179.2 (261.7) | 205.9 (279.6) |
| **Average hours of sleep** | 7.3 (1.1) | 7.3 (1.0) | 7.1 (1.3) | 7.4 (1.3) |
| **Current smoking, %** | 10.6% | 7.8% | 26.1% | 8.5% |

Data on means and percentages are averaged from 10 imputed datasets. Abbreviations. LE8: Life's Essential 8; CVH: cardiovascular health; NH: Non-Hispanic; BMI: body mass index; HEI: Healthy Eating Index.

### Contributors to racial and ethnic gaps in CVH

Differences in early pregnancy CVH, followed by SDOH, accounted for the largest share of the Black-White and Hispanic-White gaps in post-pregnancy CVH. Specifically, **Table 3** present results from the Oaxaca-Blinder decomposition for racial and ethnic differences in post-pregnancy CVH. The observed White-Black and White-Hispanic gaps in mean post-pregnancy CVH scores were 12.2 points (95% CI, 10.3-14.2) and 4.6 points (95% CI, 1.9-7.3), respectively.

**Table 3.** Oaxaca-Blinder Decomposition Quantifying Racial and Ethnic Differences in Post-pregnancy LE8 CVH Scores *Explained* by Differences in Mean Levels of early pregnancy CVH and SDOH.

|  | NH White compared with NH Black women |  | NH White compared with Hispanic women |  |
| --- | --- | --- | --- | --- |
|  | Estimate (95% CI) | % of total | Estimate (95% CI) | % of total |
| **Total group difference** | 12.2 (10.3, 14.2) |  | 4.6 (1.9, 7.3) |  |
| **Explained by differences in risk factors** | **9.0 (6.8, 11.3)** | **74%** | **3.8 (0.7, 6.8)** | **83%** |
| **Early pregnancy CVH** | **5.5 (4.2, 6.8)** | **45%** | **2.6 (0.9, 4.4)** | **57%** |
| **Demographic subtotal** | **-1.3 (-1.9, -0.6)** | **-11%** | **-2.2 (-3.3, -1.0)** | **-48%** |
| Maternal age | -1.3 (-2.0, -0.5) | -11% | -0.9 (-1.5, -0.3) | -20% |
| Nativity | -0.0 (-0.1, 0.1) | 0% | -1.3 (-2.1, -0.4) | -28% |
| **SDOH subtotal** | **4.5 (2.8, 6.2)** | **37%** | **3.2 (1.1, 5.3)** | **70%** |
| Individual SDOH subtotal | 3.1 (1.8, 4.5) | 25% | 2.3 (1.0, 3.7) | 50% |
| Education | 1.2 (0.7, 1.8) | 10% | 1.0 (0.5, 1.4) | 22% |
| Income | 0.7 (-0.0, 1.3) | 6% | 0.4 (-0.1, 0.8) | 9% |
| Insurance | 1.2 (0.5, 1.9) | 10% | 0.9 (0.1, 1.8) | 20% |
| Health literacy | 0.2 (-0.1, 0.6) | 2% | 0.2 (-0.3, 0.7) | 4% |
| Perceived racial discrimination | -0.3 (-0.5, -0.0) | -2% | -0.1 (-0.3, 0.1) | -2% |
| Neighborhood SDOH (ADI) | 1.4 (0.8, 2.0) | 11% | 0.9 (-0.0, 1.7) | 20% |
| **Psychosocial factors subtotal** | **0.3 (-0.0, 0.6)** | **2%** | **0.1 (-0.1, 0.2)** | **2%** |
| Resilience | -0.0 (-0.0, 0.0) | 0% | -0.0 (-0.1, 0.1) | 0% |
| Perceived social support | -0.0 (-0.2, 0.1) | 0% | -0.0 (-0.1, 0.1) | 0% |
| Anxiety symptoms | 0.1 (0.0, 0.2) | 1% | 0.1 (0.0, 0.1) | 2% |
| Depressive symptoms | -0.2 (-0.4, 0.0) | -2% | -0.1 (-0.3, 0.1) | -2% |
| Perceived stress | 0.4 (-0.1, 0.8) | 3% | 0.2 (-0.1, 0.4) | 4% |

Estimates are LE8 points with 95% confidence intervals. Positive estimates indicate that equalizing the factor would narrow the CVH gap; negative estimates indicate that equalizing the factor would widen it. Each row represents the contribution to the group difference in post-pregnancy CVH if the Black or Hispanic group assumed the same value of the factor as the White group. The SDOH subtotal comprises the individual, neighborhood, and social context domains and was estimated directly rather than by summing component rows. Perceived racial discrimination, a social context measure, was grouped with individual-level SDOH for estimation. n=3,368 for the NH White versus NH Black comparison and n=3,498 for the NH White versus Hispanic comparison. ADI, Area Deprivation Index; CI, confidence interval; CVH, cardiovascular health; LE8, Life's Essential 8; NH, non-Hispanic; SDOH, social determinants of health.

**Table 3** quantifies the expected reduction in the racial and ethnic gap in post-pregnancy CVH for Black and Hispanic versus White women if the distribution of demographic, early pregnancy CVH, SDOH, and psychosocial factors were equal to those of White women. If equality across all of these measures were achieved while maintaining the associations between these measures and post-pregnancy CVH scores, the Black-White gap in mean post-pregnancy CVH scores would be reduced by 9.0 points (95% CI, 6.8-11.3) or a 74% reduction of the total gap; and the Hispanic-White gap in post-pregnancy CVH would be reduced by 3.8 points (95% CI, 0.7-6.8) or 83% of the total gap.

The greatest reductions in the Black-White and Hispanic-White gaps in mean post-pregnancy CVH scores were due to differences in early pregnancy CVH scores followed by SDOH (**Table 3**). If early pregnancy CVH scores for Black and Hispanic women were the same as White women, holding everything else constant, the Black-White and Hispanic-White gaps in mean post-pregnancy CVH scores would be reduced by 5.5 points (95% CI, 4.2-6.8) or 45% of the total gap and 2.6 points (95% CI, 0.9-4.4) or 57% of the total gap, respectively. Similarly, if all early pregnancy SDOH among Black and Hispanic women were the same as White women, holding everything else constant, the Black-White and Hispanic-White gaps in mean post-pregnancy CVH scores would be reduced by 4.5 points (95% CI, 2.8-6.2) or 37% of the total gap and 3.2 points (95% CI, 1.1-5.3) or 70% of the total gap, respectively. Considered separately, individual-level SDOH, which includes perceived racial discrimination as a social context measure grouped with individual-level SDOH for estimation, accounted for 3.1 points (95% CI, 1.8-4.5) of the Black-White gap and 2.3 points (95% CI, 1.0-3.7) of the Hispanic-White gap. If the neighborhoods where Black and Hispanic women lived had the same level of socioeconomic deprivation as those where White women lived, holding everything else constant, the Black-White and Hispanic-White gaps in mean post-pregnancy CVH scores would be reduced by 1.4 points (95% CI, 0.8-2.0) or 11% of the total gap and 0.9 points (95% CI, -0.0 to 1.7) or 20% of the total gap, respectively.

The contribution of SDOH depended on whether early pregnancy CVH was included in the model (Supplemental Table S8). In Model 1, which included demographic covariates and SDOH only, differences in SDOH accounted for 10.5 points (95% CI, 7.4-13.5) of the Black-White gap, or 86% of the total difference, and 7.4 points (95% CI, 3.2-11.6) of the Hispanic-White gap. The Hispanic-White estimate exceeds the total gap of 4.6 points because the younger age and higher proportion of women born outside the US among Hispanic women contributed in the opposite direction, together accounting for -3.7 points (95% CI, -5.6 to -1.8). Adding psychosocial factors in Model 2 changed the contribution of SDOH only modestly, to 9.6 points (95% CI, 6.8-12.5) and 6.8 points (95% CI, 2.9-10.7). Adding early pregnancy CVH in Model 3, the primary model, reduced it to 4.5 points (95% CI, 2.8-6.2) and 3.2 points (95% CI, 1.1-5.3).

Some early pregnancy factors were more favorable among Black and Hispanic women compared with White women, resulting in wider racial-and ethnic-specific gaps in post-pregnancy CVH scores under hypothetical interventions to equalize them (**Table 3**). Black and Hispanic women were about 4.3 and 3.2 years younger than White women at enrollment in the nuMoM2b cohort. In the scenario where mean maternal age for Black and Hispanic women was set to mean maternal age among White women, the Black-White and Hispanic-White differences in post-pregnancy CVH scores would be 1.3 points (95% CI, 0.5 to 2.0) and 0.9 points (95% CI, 0.3 to 1.5) wider. Similarly, 43.2% of Hispanic women were born outside the US compared with 4% of White women. In the scenario where the prevalence of US nativity in Hispanic women was set to that of White women, the Hispanic-White gap in post-pregnancy CVH score would be 1.3 points (95% CI, 0.4 to 2.1) wider. Psychosocial factors contributed modestly to racial and ethnic differences in post-pregnancy CVH after adjusting for all other risk factors. In the primary model, equalizing the distributions of psychosocial factors would narrow the Black-White gap by 0.3 points (95% CI, -0.0 to 0.6) and the Hispanic-White gap by 0.1 points (95% CI, -0.1 to 0.2). Before adjustment for early pregnancy CVH, the corresponding contributions were larger and statistically significant, at 0.7 points (95% CI, 0.4-1.1) and 0.4 points (95% CI, 0.1-0.7).

### Differences in associations between SDOH and CVH (unexplained component)

Beyond differences in mean levels of risk factors, the associations between early pregnancy factors and post-pregnancy CVH differed by race and ethnicity. This unexplained, or coefficient, component captures the portion of the gap arising because equivalent levels of a given factor were associated with different post-pregnancy CVH across groups. It was 3.2 points (95% CI, 1.2-5.2), or 26% of the total difference, for the Black-White comparison, and 0.9 points (95% CI, -0.9 to 2.6), or 20%, for the Hispanic-White comparison; the latter was not statistically distinguishable from zero. The two estimates were not formally compared. Contributions of individual factors, including the intercept, are reported in Supplemental Table S10 and are imprecise and not interpreted individually.

### Change in CVH components between early pregnancy and follow-up

Overall CVH declined between early pregnancy and follow-up among Black women, by - 2.2 points (95% CI, -3.9 to -0.6), and was essentially unchanged among White women (-0.0 points; 95% CI, -1.1 to 1.0) and Hispanic women (0.2 points; 95% CI, -1.9 to 2.2) (Supplemental Tables S6 and S7). Among Black women the largest component declines were in body mass index (-8.1 points), lipids (-7.7 points), and blood pressure (-6.0 points). Physical activity and nicotine exposure scores improved among White women (9.5 and 6.9 points) and Hispanic women (15.0 and 4.0 points) but not among Black women (8.2 and -2.0 points). Lipid scores declined by approximately 8 points in all three groups. These within-group changes are descriptive and were not compared across groups.

In analyses restricted to participants with complete data (n=2,540 for the Black-White comparison and n=2,618 for the Hispanic-White comparison), the contribution of SDOH in the primary model was 4.0 points (95% CI, 2.1-5.9) and 2.1 points (95% CI, 0.4-3.8), consistent with the primary analysis (Supplemental Table S9). Residuals from the group-specific models were approximately homoscedastic and approximately normally distributed, with mild departure in the lower tail (Supplemental Figure S1).

## DISCUSSION

In a prospective US cohort of pregnant women followed from their first pregnancy through 2-to-7 years after their first delivery, post-pregnancy CVH was worse, as measured by the AHA LE8, among Black and Hispanic women compared with White women. Using Oaxaca-Blinder decomposition analysis, we found that racial and ethnic differences in post-pregnancy CVH were primarily explained by early differences in early pregnancy CVH followed by SDOH.

Racial and ethnic differences in post-pregnancy CVH observed in the current analysis extend epidemiologic associations observed in prior studies including differences in early pregnancy CVH^13^and SDOH.^43^

The smaller independent contribution of psychosocial factors in the current analysis reflects in part their position on the pathway between SDOH and CVH: their contribution was statistically significant before adjustment for early pregnancy CVH and attenuated toward the null thereafter. Among Black women, a statistically significant unexplained component was observed, indicating that the associations between measured early pregnancy factors and post-pregnancy CVH differed from those among White women. The corresponding estimate among Hispanic women was not statistically distinguishable from zero. A differential association of this kind is consistent with evidence that minoritized racial and ethnic groups may derive smaller health returns from equivalent socioeconomic resources, reflecting structural and interpersonal racism, residential segregation, differential access to and quality of health care, and the cumulative physiologic burden of chronic stress.^44,45^ That the estimate among Hispanic women was null may reflect the substantially greater heterogeneity of this group, of whom 43.2% were born outside the US compared with 5.4% of Black women, together with a smaller total difference to decompose and correspondingly less precision.

This differential association carries a direct implication for interpretation. If equivalent social and economic resources do not yield equivalent cardiovascular health, then equalizing the levels of measured social determinants would not by itself eliminate the difference in post-pregnancy CVH. In the current analysis, approximately 26% of the Black-White difference would be expected to persist even if the distributions of all measured early pregnancy factors were equalized. Interventions that target the levels of social risk factors are therefore necessary but not sufficient, and addressing the structural conditions that produce unequal returns to those resources is also required.

These findings extend a broader literature on cardiovascular health across the life course. In cohorts of young adults not selected for pregnancy, cardiovascular health measured in late adolescence and early adulthood predicts premature cardiovascular disease and mortality.^46^ Differences in cardiovascular health also account for a substantial share of Black-White differences in cardiovascular disease incidence and mortality in the general adult population,^47^ consistent with decomposition analyses of cardiovascular health in non-pregnant samples.^18^ Professional guidance has likewise identified the period after delivery as an opportunity to reduce cardiovascular risk.^48^ The current findings locate that pattern within the reproductive years and indicate that the social circumstances present at the start of a first pregnancy remain associated with cardiovascular health years after that pregnancy has ended.

The component-level findings point to a distinct target. The decline in overall CVH among Black women was driven not only by worsening body mass index and blood pressure but by the absence of the improvements in physical activity and nicotine exposure observed among White and Hispanic women over the same interval. Because Black women had lower component scores in early pregnancy, regression to the mean would be expected to move their follow-up scores upward rather than downward, so the observed decline is unlikely to be an artifact of baseline differences. Interventions in the years after a first birth that support physical activity and smoking cessation may therefore address a component of the difference that is not captured by early pregnancy risk factor levels alone.

The current study emphasizes that, independent of early pregnancy CVH, SDOH remain important determinants of long-term racial and ethnic post-pregnancy CVH disparities. These findings underscore the need for developing and testing interventions that address SDOH disparities early in pregnancy or even preconceptionally to improve long-term maternal CVH.^49^ Professional societies have recommended strategies to reduce racial and ethnic differences in periconception CVH and maternal morbidity, including community-engaged pre-pregnancy interventions, expansion of access to healthy food to address nutrition insecurity, creation of educational and employment opportunities, and promotion of collaborative peripartum care models.^4^

Several limitations warrant consideration. First, this study used an observational cohort, and the Oaxaca-Blinder decomposition quantifies statistical contributions to group differences; as such, causality cannot be known. Second, the decomposition framework is limited by the factors included in the model, and unmeasured confounders may explain additional variance in post-pregnancy CVH disparities. Third, psychosocial factors were assessed by self-report at a single early pregnancy timepoint. Their modest independent contribution should not be read as evidence that they are unimportant: they are plausibly downstream of the social determinants alongside which they are modeled, and their contribution was larger and statistically significant before adjustment for early pregnancy CVH. Single-timepoint assessment may additionally underestimate cumulative psychosocial exposure across the years between pregnancy and follow-up. Fourth, our analysis was restricted to non-Hispanic White, non-Hispanic Black, and Hispanic women, as the sample sizes of women identifying as non-Hispanic Asian, multiracial, or other racial and ethnic groups were insufficient to support stable decomposition estimates. Future studies in cohorts designed with adequate representation of these groups are needed to extend this work. Fifth, the nuMoM2b-HHS cohort comprised nulliparous individuals recruited primarily from tertiary care centers, which may limit generalizability. Sixth, LE8 component scores are bounded and, for several components, coarsely graded, so ceiling effects may attenuate observed differences among groups with more favorable scores. Finally, the analytic sample comprised participants who attended the follow-up visit; demographic characteristics were similar between included participants and nuMoM2b participants who did not enroll in the follow-up study (Supplemental Table S1), although residual differences may remain.

In conclusion, in a prospective multicenter cohort study, racial and ethnic differences in post-pregnancy CVH as measured by LE8 2-to-7 years after a first delivery were largely explained by differences in early pregnancy CVH followed by SDOH. These findings highlight the need for periconception CVH optimization and structural interventions targeting SDOH inequities to promote equitable CVH across a woman’s life course.

## Data Availability

The data that support the findings of this study are available from the Eunice Kennedy Shriver National Institute of Child Health and Human Development Data and Specimen Hub (DASH) for investigators who meet the criteria for access to de-identified data.

## Acknowledgments

The authors thank the nuMoM2b and nuMoM2b Heart Health Study investigators, study staff, and participants. The parent studies were supported by grant funding from the Eunice Kennedy Shriver National Institute of Child Health and Human Development (NICHD): U10 HD063036; U10 HD063072; U10 HD063047; U10 HD063037; U10 HD063041; U10 HD063020; U10 HD063046; U10 HD063048; and U10 HD063053. In addition, support was provided by Clinical and Translational Science Institutes: UL1TR001108 and UL1TR000153. The nuMoM2b Heart Health Study was supported by the National Heart, Lung, and Blood Institute (U10 HL119991). The content is solely the responsibility of the authors and does not necessarily represent the official views of the National Institutes of Health.

## Supplemental Material

Supplemental Table S1. Early pregnancy characteristics by sample inclusion and exclusion criteria.

Supplemental Table S2. Multiple imputation model specification for variables with missing data.

Supplemental Table S3. Early pregnancy characteristics by racial and ethnic group, before imputation.

Supplemental Table S4. Post-pregnancy CVH factors and LE8 score by racial and ethnic group, before imputation.

Supplemental Table S5. LE8 component scoring as operationalized in nuMoM2b-HHS.

Supplemental Table S6. Early pregnancy CVH factors and LE8 component scores by racial and ethnic groups.

Supplemental Table S7. Within-group change in LE8 component scores between early pregnancy and follow-up.

Supplemental Table S8. Oaxaca-Blinder decomposition with sequential addition of psychosocial factors and early pregnancy CVH.

Supplemental Table S9. Oaxaca-Blinder decomposition restricted to participants with complete data.

Supplemental Table S10. Oaxaca-Blinder decomposition of the unexplained component.

Supplemental Figure S1. Residuals plots, by racial and ethnic groups.

