## Supplemental Materials for "Impact of Social Determinants of Health in Early Pregnancy on Racial and Ethnic Differences in Post-pregnancy Cardiovascular Health"

**Supplemental Material**

Supplemental Table S1. Early pregnancy characteristics by sample inclusion and exclusion criteria.

Supplemental Table S2. Multiple imputation model specification for variables with missing data.

Supplemental Table S3. Early pregnancy characteristics by racial and ethnic group, before imputation.

Supplemental Table S4. Post-pregnancy CVH factors and LE8 score by racial and ethnic group, before imputation.

Supplemental Table S5. LE8 component scoring as operationalized in nuMoM2b-HHS.

Supplemental Table S6. Early pregnancy CVH factors and LE8 component scores by racial and ethnic groups.

Supplemental Table S7. Within-group change in LE8 component scores between early pregnancy and follow-up.

Supplemental Table S8. Oaxaca-Blinder decomposition with sequential addition of psychosocial factors and early pregnancy CVH.

Supplemental Table S9. Oaxaca-Blinder decomposition restricted to participants with complete data.

Supplemental Table S10. Oaxaca-Blinder decomposition of the unexplained component.

Supplemental Figure S1. Residuals plots, by racial and ethnic groups.

**Supplemental Table S1. Early pregnancy Demographic, SDOH, and Psychosocial Characteristics, and LE8 CVH by sample inclusion/exclusion criteria**

|  | **Analytical sample** | **Excluded: Other race/ethnicity** | **Excluded: Age<18** | **Excluded: Non-HHS1 participants** | **Total** |
| --- | --- | --- | --- | --- | --- |
|  | **N=4,088** | **N=931** | **N=214** | **N=4,805** | **N=10,038** |
| **Demographic characteristics** |  |  |  |  |  |
| **Maternal Age, years, mean (SD)** | 27.1 (5.4) | 28.0 (6.1) | 16.4 (0.8) | 27.1 (5.5) | 26.9 (5.7) |
| **Nativity** |  |  |  |  |  |
| **US-born, %** | 88.9% | 63.2% | 91.1% | 88.8% | 86.5% |
| **Born outside the US, %** | 11.1% | 36.8% | 8.9% | 11.2% | 13.5% |
| **Gestational age in weeks at screening, mean (SD)** | 11.4 (1.6) | 11.6 (1.5) | 11.6 (1.6) | 11.6 (1.5) | 11.5 (1.5) |
| **SDOH characteristics** |  |  |  |  |  |
| **Education (years of schooling), mean (SD)** | 15.2 (2.4) | 15.6 (2.8) | 10.2 (1.0) | 15.2 (2.6) | 15.2 (2.6) |
| **Household income (per $10,000), mean (SD)** | 8.4 (5.8) | 9.4 (6.3) | 2.7 (2.5) | 9.1 (6.2) | 8.7 (6.1) |
| **Health insurance** |  |  |  |  |  |
| **Others, %** | 3.3% | 5.4% | 1.9% | 4.0% | 3.8% |
| **Covered by public insurance, %** | 26.6% | 26.3% | 76.0% | 27.7% | 28.1% |
| **Covered by private insurance, %** | 70.1% | 68.3% | 22.1% | 68.3% | 68.1% |
| **Adequate health literacy, %** | 83.8% | 81.6% | 40.5% | 83.4% | 82.5% |
| **Area deprivation index 2015, national rank, mean (SD)** | 47.5 (30.3) | 42.0 (31.6) | 67.8 (27.5) | 42.9 (30.4) | 45.2 (30.7) |
| **Psychosocial factors** |  |  |  |  |  |
| **Connor-Davidson Resilience Scale (0-100), mean (SD)** | 79.1 (12.2) | 77.8 (12.4) | 72.1 (15.9) | 79.3 (12.1) | 78.9 (12.3) |
| **Multidimensional Scale of Perceived Social Support (0-72), mean (SD)** | 62.6 (14.0) | 61.6 (13.8) | 56.1 (13.9) | 62.4 (14.3) | 62.3 (14.2) |
| **State-Trait Anxiety Inventory (0-60), mean (SD)** | 13.6 (8.7) | 14.3 (8.4) | 18.8 (8.7) | 13.8 (8.6) | 13.9 (8.7) |
| **Edinburgh Postnatal Depression Scale (0-30), mean (SD)** | 5.7 (4.2) | 6.1 (4.1) | 7.3 (4.9) | 5.8 (4.2) | 5.8 (4.2) |
| **PSS-10 stress scale (0-40), mean (SD)** | 12.5 (6.7) | 13.6 (6.5) | 17.1 (7.1) | 12.7 (6.6) | 12.8 (6.7) |
| **Experienced racial discrimination, %** | 5.8% | 15.0% | 5.0% | 6.1% | 6.8% |
| **LE8 CVH Score, mean (SD)** | 78.5 (13.7) | 78.0 (15.6) | 73.8 (14.4) | 74.9 (16.5) | 76.6 (15.4) |

LE8: Life’s Essential 8; CVH: cardiovascular health; NH: Non-Hispanic; SD: standard deviation

**Supplemental Table S2. Multiple imputation model specification for variables with missing data**

| **Variable** | **% Missing** | **Imputation Model** |
| --- | --- | --- |
| **Demographic and SES** |  |  |
| Health insurance | .6 | Multinomial logistic regression |
| Household income in 10K USD | 17.8 | PMM, k=10 |
| Nativity | .4 | Logistic regression |
| Health literacy | 4.0 | Logistic regression |
| Experienced racial discrimination | 3.1 | Logistic regression |
| Connor-Davidson Resilience Scale | 3.9 | PMM, k=10 |
| Multidimensional Scale of Perceived Social Support | 10.5 | PMM, k=10 |
| State-Trait Anxiety Inventory | 10.4 | PMM, k=10 |
| Edinburgh Postnatal Depression Scale | 2.4 | PMM, k=10 |
| PSS-10 stress scale | .4 | PMM, k=10 |
| Area deprivation index 2015, national rank | 4.4 | PMM, k=10 |
| **Early pregnancy CVH** |  |  |
| HEI 2010 Total Score | 14.8 | PMM, k=10 |
| Moderate-to-vigorous physical activity | 4.6 | PMM, k=10 |
| Current smoking | .1 | PMM, k=10 |
| Average hours of sleep | 21.2 | PMM, k=10 |
| BMI | 1.8 | PMM, k=10 |
| Non-HDL-cholesterol | 2.5 | PMM, k=10 |
| Glucose | 2.5 | PMM, k=10 |
| SBP | 2.3 | PMM, k=10 |
| DBP | 2.3 | PMM, k=10 |
| **Post-pregnancy CVH** |  |  |
| HEI 2010 Total Score | 20.4 | PMM, k=10 |
| Moderate-to-vigorous physical activity | 3.7 | PMM, k=10 |
| Current smoking | .1 | PMM, k=10 |
| Average hours of sleep | 18.4 | PMM, k=10 |
| BMI | .5 | PMM, k=10 |
| Non-HDL-cholesterol | 1.9 | PMM, k=10 |
| Glucose | 1.9 | PMM, k=10 |
| SBP | .2 | PMM, k=10 |
| DBP | .2 | PMM, k=10 |

Multiple imputation by chained equations to generate 10 datasets. Non-missing variables include maternal age, gestational age at screening, race/ethnicity, study sites, medication use for hypertension, cholesterol, or diabetes. PMM: predictive mean matching.

**Supplemental Table S3. Early pregnancy Demographic, SDOH, and Psychosocial Characteristics, and LE8 CVH Overall and by Racial and Ethnic Group (before imputation, nuMoM2b-HHS, N=4,088)**

|  | **Total** | **NH White** | **NH Black** | **Hispanic** |
| --- | --- | --- | --- | --- |
|  | **N=4,088** | **N=2,778** | **N=590** | **N=720** |
| **Demographic characteristics** |  |  |  |  |
| **Maternal Age, years, mean (SD)** | 27.1 (5.4) | 28.3 (5.1) | 24.0 (5.2) | 25.1 (5.3) |
| **Nativity** |  |  |  |  |
| **US-born, %** | 88.9% | 96.0% | 94.6% | 56.8% |
| **Born outside the US, %** | 11.1% | 4.0% | 5.4% | 43.2% |
| **Gestational age in weeks at screening, mean (SD)** | 11.4 (1.6) | 11.4 (1.6) | 11.5 (1.6) | 11.5 (1.5) |
| **Individual SDOH characteristics** |  |  |  |  |
| **Education (years of schooling), mean (SD)** | 15.2 (2.4) | 15.9 (2.2) | 13.5 (2.2) | 14.2 (2.4) |
| **Household income (per $10,000), mean (SD)** | 8.4 (5.8) | 9.4 (5.7) | 4.1 (4.5) | 6.0 (5.2) |
| **Health insurance** |  |  |  |  |
| **Others, %** | 3.3% | 2.9% | 4.5% | 4.2% |
| **Covered by public insurance, %** | 26.6% | 12.4% | 58.8% | 55.2% |
| **Covered by private insurance, %** | 70.1% | 84.7% | 36.7% | 40.6% |
| **Adequate health literacy, %** | 83.8% | 92.6% | 64.5% | 62.4% |
| **Area deprivation index 2015, national rank, mean (SD)** | 47.5 (30.3) | 38.9 (25.4) | 73.8 (25.8) | 60.7 (34.4) |
| **Psychosocial factors** |  |  |  |  |
| **Connor-Davidson Resilience Scale (0-100), mean (SD)** | 79.1 (12.2) | 79.6 (11.3) | 78.8 (14.4) | 77.2 (13.6) |
| **Multidimensional Scale of Perceived Social Support (0-72), mean (SD)** | 62.6 (14.0) | 63.6 (13.3) | 57.8 (16.5) | 61.3 (13.9) |
| **State-Trait Anxiety Inventory (0-60), mean (SD)** | 13.6 (8.7) | 13.3 (8.6) | 15.1 (9.2) | 14.2 (9.0) |
| **Edinburgh Postnatal Depression Scale (0-30), mean (SD)** | 5.7 (4.2) | 5.4 (4.0) | 6.4 (4.8) | 6.2 (4.4) |
| **PSS-10 stress scale (0-40), mean (SD)** | 12.5 (6.7) | 11.8 (6.3) | 14.9 (7.3) | 13.6 (7.1) |
| **Experienced racial discrimination, %** | 5.8% | 1.9% | 18.8% | 10.8% |
| **LE8 CVH Score, mean (SD)** | 80.0 (13.9) | 82.4 (13.2) | 72.2 (14.2) | 77.2 (13.0) |

LE8: Life’s Essential 8; CVH: cardiovascular health; NH: Non-Hispanic; SD: standard deviation; SDOH: social determinants of health.

**Supplemental Table S4. Post-pregnancy Cardiovascular Health Factors and Composite LE8 CVH Score 2 to 7 Years After Delivery, Overall and by Racial and Ethnic Group (before imputation, nuMoM2b-HHS, N=4,088)**

|  | **Total** | **NH White** | **NH Black** | **Hispanic** |
| --- | --- | --- | --- | --- |
|  | **N=4,088** | **N=2,778** | **N=590** | **N=720** |
| **Life’s Essential 8 Scores** |  |  |  |  |
| **Blood Pressure Score** | 85.8 (24.8) | 86.8 (23.5) | 79.2 (29.6) | 87.2 (24.2) |
| **Lipid Score** | 78.7 (27.4) | 77.5 (28.0) | 84.4 (24.4) | 78.8 (26.8) |
| **Glucose Score** | 91.1 (22.6) | 92.4 (20.6) | 86.7 (28.6) | 89.5 (24.1) |
| **BMI Score** | 67.5 (35.8) | 72.7 (33.8) | 48.0 (38.3) | 63.2 (35.3) |
| **Diet Score** | 71.6 (24.2) | 74.3 (22.4) | 56.0 (29.4) | 70.9 (23.8) |
| **Physical Activity Score** | 68.0 (40.7) | 74.4 (37.3) | 51.7 (43.9) | 55.8 (44.3) |
| **Sleep Health Score** | 87.3 (20.5) | 89.5 (18.4) | 79.7 (24.9) | 84.8 (22.5) |
| **Nicotine Exposure Score** | 89.4 (30.7) | 92.2 (26.8) | 73.9 (44.0) | 91.5 (27.9) |
| **Overall CVH** | 79.9 (14.1) | 82.5 (13.1) | 70.6 (14.9) | 77.7 (13.4) |
| **CVH risk factors** |  |  |  |  |
| **Mean Arterial Pressure (mmHg)** | 85.3 (9.8) | 85.0 (9.4) | 87.4 (11.6) | 84.8 (9.3) |
| **Non-HDL-cholesterol (mg/dL)** | 126.1 (37.4) | 128.3 (38.0) | 116.0 (35.1) | 125.6 (35.6) |
| **Glucose (mg/dL)** | 90.9 (21.0) | 89.7 (17.1) | 95.3 (35.3) | 92.1 (18.3) |
| **BMI (kg/m^2)** | 27.9 (7.7) | 26.8 (7.1) | 32.2 (9.2) | 28.8 (7.4) |
| **HEI 2010 Total Score** | 62.7 (11.7) | 64.0 (11.3) | 55.3 (11.8) | 62.0 (11.5) |
| **Physical activity (min/week)** | 248.9 (279.4) | 275.0 (278.9) | 174.2 (259.2) | 206.2 (280.8) |
| **Average hours of sleep** | 7.2 (1.1) | 7.3 (1.0) | 7.1 (1.4) | 7.4 (1.3) |
| **Current smoking** | 10.6% | 7.8% | 26.1% | 8.5% |

Data are presented as mean (SD) or percentage. LE8: Life’s Essential 8; CVH: cardiovascular health; NH: Non-Hispanic; BMI: body mass index; HEI: Healthy Eating Index.

**Supplemental Table S5. Life’s Essential 8 component scoring as operationalized in nuMoM2b-HHS**

| **Component** | **Measure and source** | **Scoring** |
| --- | --- | --- |
| Blood pressure | Measured by trained personnel using a standardized protocol | 100: <120/<80; 75: 120-129/<80; 50: 130-139 or 80-89; 25: 140-159 or 90-99; 0: 160+ or 100+. Subtract 20 if treated |
| Body mass index | Measured height and weight | 100: <25.0; 70: 25.0-29.9; 30: 30.0-34.9; 15: 35.0-39.9; 0: 40.0+ |
| Non-HDL cholesterol | Non-fasting sample at visit 1; fasting at follow-up | 100: <130; 60: 130-159; 40: 160-189; 20: 190-219; 0: 220+ mg/dL. Subtract 20 if drug-treated |
| Blood glucose | Non-fasting sample at visit 1; fasting at follow-up | 100: <100; 60: 100 to <126; 0: 126+ mg/dL or treated for / reporting diabetes |
| Diet | HEI-2010 from the Block/Bodnar/nuMoM2b 2010 food frequency questionnaire; reference period 3 months pre-conception at visit 1 and past year at follow-up | Percentile of the HEI-2010 population distribution. 100: above 95th; 80: 75th-94th; 50: 50th-74th; 25: 25th-49th; 0: 1st-24th |
| Physical activity | Modifiable Activity Questionnaire; leisure-time moderate-to-vigorous activity using MET values from the 2011 Compendium, excluding occupational and household activity and activity below 3.0 METs, vigorous minutes weighted twice | Minutes per week. 100: 150+; 90: 120-149; 80: 90-119; 60: 60-89; 40: 30-59; 20: 1-29; 0: 0 |
| Sleep health | Self-reported average hours per night | 100: 7 to <9; 90: 9 to <10; 70: 6 to <7; 40: 5 to <6 or 10+; 20: 4 to <5; 0: <4 |
| Nicotine exposure | Self-reported smoking status | 100: never smoker; 50: former smoker; 0: current smoker |

The composite LE8 score is the individual-specific mean of the eight component scores, ranging from 0 to 100.

**Supplemental Table S6. Early pregnancy cardiovascular health factors and LE8 component scores, overall and by racial and ethnic group**

|  | **Total** | **NH White** | **NH Black** | **Hispanic** |
| --- | --- | --- | --- | --- |
|  | **N=4,088** | **N=2,778** | **N=590** | **N=720** |
| **Mean Arterial Pressure (mmHg)** | 81.5 (8.2) | 81.6 (8.0) | 83.0 (8.8) | 80.2 (8.0) |
| **Non-HDL-cholesterol (mg/dL)** | 114.5 (30.9) | 116.9 (31.4) | 104.3 (27.8) | 113.5 (29.5) |
| **Glucose (mg/dL)** | 87.9 (15.7) | 87.6 (15.0) | 88.1 (18.0) | 89.0 (16.4) |
| **BMI (kg/m^2)** | 26.7 (6.5) | 25.9 (5.9) | 30.2 (8.5) | 26.9 (6.0) |
| **HEI 2010 Total Score** | 62.5 (12.7) | 64.9 (12.2) | 53.7 (11.0) | 60.5 (12.3) |
| **Moderate-to-vigorous physical activity (min/week)** | 163.3 (217.6) | 184.5 (220.1) | 124.7 (213.8) | 113.3 (197.6) |
| **Average hours of sleep** | 8.0 (1.3) | 7.9 (1.1) | 7.9 (1.6) | 8.1 (1.4) |
| **Current smoking** | 15.6% | 14.7% | 24.1% | 12.5% |
| **Blood Pressure Score** | 90.0 (19.1) | 90.3 (18.5) | 85.2 (22.8) | 92.5 (17.0) |
| **Lipid Score** | 86.8 (22.2) | 85.5 (23.1) | 92.1 (17.0) | 87.4 (21.4) |
| **Glucose Score** | 89.7 (24.1) | 90.7 (22.8) | 88.1 (27.3) | 87.4 (26.0) |
| **BMI Score** | 73.0 (33.2) | 77.1 (31.2) | 56.1 (38.3) | 71.4 (31.9) |
| **Diet Score** | 70.5 (26.7) | 74.9 (24.4) | 53.1 (29.4) | 67.9 (26.4) |
| **Physical Activity Score** | 57.7 (43.8) | 64.9 (41.6) | 44.5 (43.7) | 40.9 (44.9) |
| **Sleep Health Score** | 88.6 (20.2) | 90.4 (18.4) | 83.8 (23.3) | 85.8 (22.7) |
| **Nicotine Exposure Score** | 84.4 (36.3) | 85.3 (35.4) | 75.9 (42.8) | 87.5 (33.1) |
| **Overall LE8 CVH Score** | 80.1 (13.2) | 82.4 (12.9) | 72.4 (12.9) | 77.6 (11.8) |

Data are mean (SD) or percentage, combined across 10 imputed datasets using Rubin’s rules. Measures were obtained at the first study visit at a mean gestational age of 11.4 weeks, except dietary quality, for which the food frequency questionnaire reference period was the 3 months before conception. Blood pressure, lipid, and glucose values in early pregnancy reflect first trimester physiologic adaptation and are not directly comparable with post-pregnancy values. Lipid and glucose values in early pregnancy were obtained from non-fasting samples; follow-up values were obtained from fasting samples. BMI, body mass index; HEI, Healthy Eating Index; LE8, Life’s Essential 8; NH, non-Hispanic; SD, standard deviation.

**Supplemental Table S7. Change in LE8 component scores between early pregnancy and follow-up, by racial and ethnic group**

|  | **NH White** | **NH Black** | **Hispanic** |
| --- | --- | --- | --- |
|  | **N=2,778** | **N=590** | **N=720** |
| Blood Pressure Score | -3.5 (-6.8, -0.3) | -6.0 (-9.1, -2.9) | -5.3 (-9.8, -0.8) |
| Lipid Score | -8.0 (-11.7, -4.3) | -7.7 (-12.0, -3.5) | -8.7 (-12.0, -5.4) |
| Glucose Score | 1.8 (-0.6, 4.1) | -1.1 (-5.2, 2.9) | 2.2 (1.0, 3.4) |
| BMI Score | -4.3 (-6.3, -2.2) | -8.1 (-10.1, -6.2) | -8.2 (-11.7, -4.7) |
| Diet Score | -1.1 (-3.7, 1.4) | 1.9 (-3.8, 7.6) | 2.4 (0.5, 4.3) |
| Physical Activity Score | 9.5 (2.9, 16.0) | 8.2 (-1.0, 17.5) | 15.0 (0.0, 30.0) |
| Sleep Health Score | -1.2 (-4.0, 1.5) | -2.9 (-8.2, 2.3) | -0.3 (-2.8, 2.3) |
| Nicotine Exposure Score | 6.9 (3.2, 10.6) | -2.0 (-6.6, 2.5) | 4.0 (0.4, 7.7) |
| Overall LE8 CVH Score | -0.0 (-1.1, 1.0) | -2.2 (-3.9, -0.6) | 0.2 (-1.9, 2.2) |

Values are the within-group mean change in LE8 score from early pregnancy to the follow-up visit 2 to 7 years after delivery, with 95% confidence intervals, estimated across 10 imputed datasets with standard errors clustered on study site. Positive values indicate improvement. The dietary instrument referred to the 3 months before conception in early pregnancy and to the past year at follow-up. Blood pressure and lipid changes partly reflect first trimester physiologic adaptation and, for lipids and glucose, the difference between non-fasting sampling in early pregnancy and fasting sampling at follow-up. LE8 component scores are bounded at 0 and 100.

**Supplemental Table S8. Oaxaca-Blinder decomposition of racial and ethnic differences in post-pregnancy LE8 CVH score, with sequential addition of psychosocial factors and early pregnancy CVH**

|  | **NH White compared with NH Black Individuals** | | | **NH White compared with Hispanic Individuals** | | |
| --- | --- | --- | --- | --- | --- | --- |
|  | **Model 1** | **Model 2** | **Model 3** | **Model 1** | **Model 2** | **Model 3** |
|  | **Est. (95% CI)** | **Est. (95% CI)** | **Est. (95% CI)** | **Est. (95% CI)** | **Est. (95% CI)** | **Est. (95% CI)** |
| Total group difference | 12.2 (10.1, 14.4) | 12.2 (10.0, 14.5) | 12.2 (10.3, 14.2) | 4.6 (2.4, 6.9) | 4.6 (2.4, 6.8) | 4.6 (1.9, 7.3) |
| Explained | 8.2 (5.7, 10.7) | 8.2 (5.8, 10.6) | 9.0 (6.8, 11.3) | 3.7 (0.5, 6.9) | 3.7 (0.6, 6.7) | 3.8 (0.7, 6.8) |
| Unexplained | 4.0 (1.0, 7.0) | 4.0 (1.0, 7.1) | 3.2 (1.2, 5.2) | 0.9 (-1.9, 3.7) | 0.9 (-1.6, 3.5) | 0.9 (-0.9, 2.6) |
| Demographic subtotal | -2.3 (-3.3, -1.2) | -2.1 (-3.1, -1.1) | -1.3 (-1.9, -0.6) | -3.7 (-5.6, -1.8) | -3.5 (-5.3, -1.8) | -2.2 (-3.3, -1.0) |
| SDOH subtotal | 10.5 (7.4, 13.5) | 9.6 (6.8, 12.5) | 4.5 (2.8, 6.2) | 7.4 (3.2, 11.6) | 6.8 (2.9, 10.7) | 3.2 (1.1, 5.3) |
| Individual SDOH | 7.9 (5.6, 10.2) | 7.1 (5.0, 9.3) | 3.1 (1.8, 4.5) | 5.9 (3.1, 8.8) | 5.4 (2.8, 8.0) | 2.3 (1.0, 3.7) |
| Neighborhood SDOH | 2.6 (1.2, 3.9) | 2.5 (1.2, 3.7) | 1.4 (0.8, 2.0) | 1.5 (-0.1, 3.0) | 1.4 (-0.0, 2.9) | 0.9 (-0.0, 1.7) |
| Psychosocial factors |  | 0.7 (0.4, 1.1) | 0.3 (-0.0, 0.6) |  | 0.4 (0.1, 0.7) | 0.1 (-0.1, 0.2) |
| Early pregnancy CVH |  |  | 5.5 (4.2, 6.8) |  |  | 2.6 (0.9, 4.4) |

Estimates are LE8 points with 95% confidence intervals. Model 1 includes demographic covariates and SDOH. Model 2 adds psychosocial factors. Model 3, the primary model, adds early pregnancy CVH. Comparison of Models 1 and 2 quantifies the share of the SDOH contribution operating through measured psychosocial factors. Model 3 estimates the contribution of SDOH that does not operate through CVH already present in early pregnancy. n=3,368 and n=3,498 respectively.

**Supplemental Table S9. Oaxaca-Blinder decomposition restricted to participants with complete data**

|  | **NH White compared with NH Black Individuals** | | | **NH White compared with Hispanic Individuals** | | |
| --- | --- | --- | --- | --- | --- | --- |
|  | **Model 1** | **Model 2** | **Model 3** | **Model 1** | **Model 2** | **Model 3** |
|  | **Est. (95% CI)** | **Est. (95% CI)** | **Est. (95% CI)** | **Est. (95% CI)** | **Est. (95% CI)** | **Est. (95% CI)** |
| Total group difference | 11.4 (9.4, 13.4) | 10.8 (8.1, 13.5) | 10.8 (8.7, 12.9) | 4.4 (1.9, 7.0) | 4.5 (2.2, 6.9) | 4.5 (1.6, 7.5) |
| Explained | 7.3 (4.4, 10.1) | 6.9 (4.0, 9.7) | 8.1 (6.3, 9.9) | 2.6 (0.4, 4.8) | 2.4 (0.2, 4.6) | 3.3 (0.5, 6.1) |
| Unexplained | 4.1 (0.5, 7.7) | 3.9 (-0.2, 8.0) | 2.7 (0.0, 5.3) | 1.8 (-0.7, 4.4) | 2.1 (0.1, 4.1) | 1.2 (-0.1, 2.6) |
| Demographic subtotal | -2.1 (-3.5, -0.8) | -1.9 (-3.2, -0.6) | -1.2 (-2.1, -0.4) | -2.2 (-3.4, -1.0) | -2.2 (-3.3, -1.0) | -1.4 (-2.2, -0.6) |
| SDOH subtotal | 9.4 (5.8, 13.0) | 8.2 (4.8, 11.6) | 4.0 (2.1, 5.9) | 4.8 (1.8, 7.7) | 4.2 (1.4, 7.0) | 2.1 (0.4, 3.8) |
| Individual SDOH | 7.0 (4.2, 9.8) | 5.9 (3.3, 8.5) | 2.6 (1.0, 4.2) | 3.8 (1.8, 5.9) | 3.4 (1.4, 5.4) | 1.6 (0.3, 2.9) |
| Neighborhood SDOH | 2.4 (1.1, 3.8) | 2.3 (1.0, 3.6) | 1.4 (0.9, 1.9) | 0.9 (-0.2, 2.0) | 0.8 (-0.2, 1.8) | 0.5 (-0.1, 1.0) |
| Psychosocial factors |  | 0.6 (0.1, 1.0) | 0.2 (-0.1, 0.5) |  | 0.3 (-0.1, 0.8) | 0.1 (-0.1, 0.3) |
| Early pregnancy CVH |  |  | 5.1 (4.2, 6.0) |  |  | 2.6 (0.7, 4.4) |
| Number of Observations | 2763 | 2540 | 2540 | 2804 | 2618 | 2618 |

Estimates are LE8 points with 95% confidence intervals. Model 1 includes demographic covariates and SDOH. Model 2 adds psychosocial factors. Model 3, the primary model, adds early pregnancy CVH. Comparison of Models 1 and 2 quantifies the share of the SDOH contribution operating through measured psychosocial factors. Model 3 estimates the contribution of SDOH that does not operate through CVH already present in early pregnancy. Analyses restricted to participants with non-missing data on all variables in the corresponding model.

**Supplemental Table S10. Oaxaca-Blinder decomposition of the unexplained component, primary model**

|  | **NH White compared with NH Black Individuals** | **NH White compared with Hispanic Individuals** |
| --- | --- | --- |
|  | **Estimate (95% CI)** | **Estimate (95% CI)** |
| **Total group difference** | 12.2 (10.3, 14.2) | 4.6 (1.9, 7.3) |
| **Unexplained** | 3.2 (1.2, 5.2) | 0.9 (-0.9, 2.6) |
| Early pregnancy CVH | -0.0 (-5.5, 5.5) | 2.3 (-3.4, 8.0) |
| Demographic subtotal | 2.6 (-3.5, 8.7) | 1.1 (-5.0, 7.2) |
| Maternal age | 2.6 (-3.4, 8.7) | 1.2 (-5.2, 7.6) |
| Nativity | -0.0 (-0.2, 0.1) | -0.1 (-0.7, 0.5) |
| SDOH subtotal | -6.8 (-12.6, -1.0) | -5.6 (-13.9, 2.8) |
| Education | -3.0 (-8.1, 2.0) | -5.3 (-13.4, 2.9) |
| Income | -1.0 (-2.5, 0.5) | 0.8 (-0.3, 2.0) |
| Insurance | -0.1 (-1.5, 1.4) | 0.4 (-1.5, 2.3) |
| Health literacy | -0.3 (-1.9, 1.2) | -0.4 (-1.8, 0.9) |
| Neighborhood (ADI) | -2.2 (-4.7, 0.2) | -1.0 (-2.7, 0.8) |
| Perceived racial discrimination | -0.2 (-0.5, 0.1) | -0.1 (-0.3, 0.1) |
| Psychosocial factors subtotal | 3.5 (-6.9, 14.0) | 5.3 (-6.0, 16.7) |
| Resilience | 0.4 (-5.6, 6.3) | 3.9 (-4.8, 12.6) |
| Perceived social support | 1.8 (-2.6, 6.1) | 0.9 (-3.2, 4.9) |
| Anxiety symptoms | 0.4 (-2.4, 3.3) | 0.1 (-2.0, 2.2) |
| Depressive symptoms | 0.2 (-2.3, 2.8) | 0.9 (-0.3, 2.2) |
| Perceived stress | 0.7 (-2.9, 4.4) | -0.4 (-4.4, 3.5) |
| Constant | 3.9 (-9.7, 17.5) | -2.4 (-11.2, 6.4) |

The unexplained component represents the portion of the between-group difference attributable to differences in the associations of measured early pregnancy factors with post-pregnancy CVH across racial and ethnic groups, assuming equal mean levels of those factors. The constant represents the difference in intercepts, that is, the portion not attributable to any measured factor. Results were estimated in each of the 10 imputed datasets and pooled via Rubin’s Rules. LE8: Life’s Essential 8; CVH: cardiovascular health; SDOH: social determinants of health; ADI: Area Deprivation Index; NH: Non-Hispanic; CI: confidence interval.

**Supplemental Figure S1. Residuals against fitted values and normal quantile plots of residuals, by racial and ethnic group**

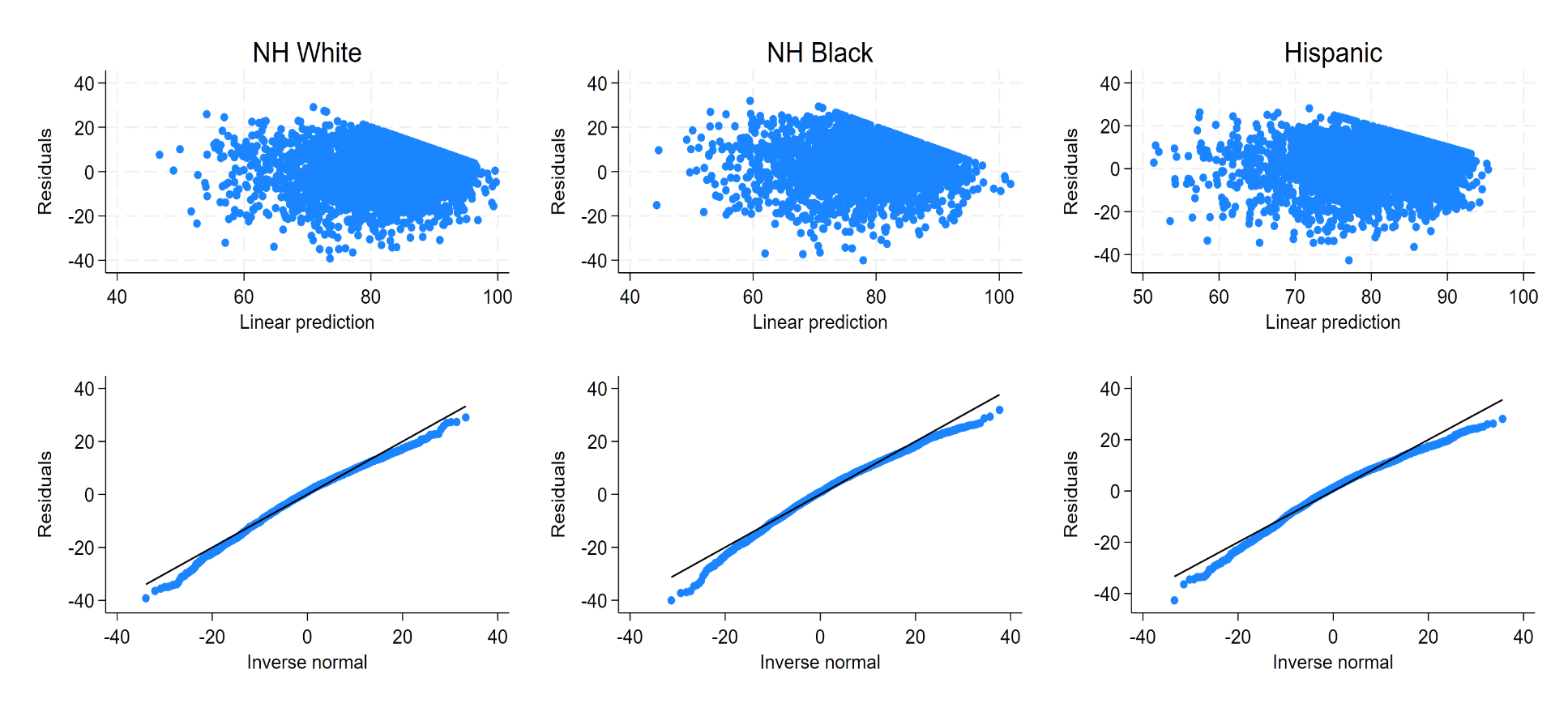

Residual and normal quantile plots: Upper panels plot residuals against fitted values from the group-specific linear models underlying the decomposition; lower panels plot residuals against normal quantiles. Residuals were approximately homoscedastic and approximately normally distributed in all three groups, with mild departure in the lower tail. The diagonal upper boundary visible in each residual plot reflects the ceiling of the LE8 score at 100, which constrains the residual to be no greater than 100 minus the fitted value.
